# MRI reveals the hierarchical organization of abdominal biological aging from shared burden to disease-specific organ engagement

**DOI:** 10.64898/2026.05.08.26352767

**Authors:** Yufeng Wang, Zengtian Deng, Lixia Wang, Abdelrahman M. Attia, Minsun Kwak, Yifan Gao, Mohammad Saeid Rezaee-Zavareh, Hyunseok Kim, Stephen J. Pandol, Sara E. Espinoza, Nicolas Musi, Judong Yang, Debiao Li

**Author notes:** Correspondence: Judong Yang and Debiao Li.

## Abstract

Multi-organ biological aging is often represented as parallel organ-specific clocks, but how age gaps should be interpreted within an anatomically coupled imaging system remains unclear. Applying end-to-end deep learning to abdominal Dixon MRI from 67,130 UK Biobank participants, we show that abdominal biological aging is hierarchically organized across eight compartments. Compartment age gaps were positively intercorrelated (mean pairwise r = 0.42), and their unweighted mean—the Overall Aging Gap (OAG)—broadly stratified all 15 prespecified prospective endpoints, including 14 incident diseases and all-cause mortality (hazard ratios 1.15–1.49 per s.d.; mortality HR = 1.41 per s.d.). After accounting for OAG, compartment-level associations became sparser and more anatomically coherent, indicating disease-specific refinement beyond the shared axis. Healthier lifestyle was associated with lower risk within accelerated-aging strata. These findings establish a hierarchical framework for interpreting abdominal MRI age gaps: OAG stratifies broad prospective risk, whereas axis-conditional compartment engagement refines disease-specific anatomical vulnerability.

## Introduction

Aging is a heterogeneous, multisystem process in which different organs within the same individual decline at different rates, and this inter-organ variation carries disease-relevant information beyond chronological age alone [1–3]. Across blood biomarkers [1], plasma proteomics [2,15], imaging-derived phenotypes [3,4] and end-to-end deep learning [5,6,8], multi-organ biological aging clocks have established that organ-level aging signals can be estimated at scale and that they stratify risk for diverse diseases and mortality. What remains less clear is how multiple organ age gaps should be interpreted together. A composite measure primarily captures between-individual differences in overall burden, while compartment-level readouts preserve anatomical resolution; whether organs and tissues within a coupled anatomical system are organized hierarchically—around a dominant shared aging axis with disease-specific compartment-level deviations conditional on that axis—has not been formally evaluated. Whether such a hierarchical readout can be further contextualized by modifiable factors such as lifestyle also remains underexplored.

The abdomen offers a particularly tractable setting for this question. A single abdominal Dixon MRI examination simultaneously captures organs and tissues central to metabolic homeostasis, body composition and systemic disease risk [9,10], and aging across these compartments is unlikely to be adequately summarized by any single organ clock or, conversely, to be entirely uncorrelated across compartments. Prior abdominal MRI aging work has already suggested that abdominal age signals are not strictly organ-autonomous: in a two-dimensional liver- and pancreas-MRI study, liver-based and pancreas-based accelerated abdominal aging were phenotypically correlated, and attention maps indicated that age prediction was driven not only by the target organs but also by surrounding abdominal organs and tissues [8]. These observations motivate a broader interpretation question: when multiple age gaps are derived from an anatomically coupled abdominal acquisition, should they be read as independent organ clocks, or as components of a coordinated abdominal aging subsystem? Recent multi-organ MRI clocks have expanded biological age estimation to additional organs and body regions [3–6], but have not directly addressed this interpretation question within a single anatomically coherent abdominal acquisition. Three-dimensional end-to-end modeling of abdominal Dixon MRI across multiple compartments may therefore better capture integrated aging variation across abdominal organs and tissues. Because these organs and body-composition tissues are exposed to shared but non-identical metabolic, hemodynamic and inflammatory stresses, aging in this region may be organized around a coordinated shared component reflecting overall abdominal burden, with compartment-specific deviations that differ across organs and tissues. The UK Biobank Imaging Study [11], with standardized abdominal Dixon MRI and linked health and lifestyle data in more than 60,000 participants, provides an opportunity to test this at scale.

Here, applying end-to-end deep learning to three-dimensional abdominal Dixon MRI from 67,130 UK Biobank participants, we estimated compartment-specific biological age across eight compartments—liver, pancreas, left kidney, right kidney, spleen, visceral adipose tissue, subcutaneous adipose tissue and skeletal muscle. Rather than treating these compartment-specific age gaps as parallel organ clocks, we asked whether they are organized hierarchically: around a dominant shared axis carrying broad prognostic information, with compartment-specific deviations conditional on that axis providing disease-specific anatomical refinement, and with lifestyle further contextualizing risk within accelerated-aging strata. To this end, we defined the Overall Aging Gap (OAG) as an unweighted summary of coordinated abdominal aging burden, operationalized axis-conditional compartment engagement using leave-one-out joint Cox models that tested each compartment after accounting for the shared-axis signal carried by the remaining compartments, and tested whether risk within OAG-defined strata was further stratified by a healthy lifestyle score. Throughout, we use ‘hierarchical’ to describe the statistical organization of abdominal age gaps—a dominant shared axis with axis-conditional compartment-specific deviations—rather than a causal hierarchy among organs.

## Results

### An integrated abdominal aging framework

We estimated biological age across eight abdominal compartments—liver, pancreas, left kidney, right kidney, spleen, visceral adipose tissue (VAT), subcutaneous adipose tissue (SAT) and skeletal muscle—by training independent deep learning models (MedNeXt) on three-dimensional Dixon MRI from a strictly healthy UK Biobank subcohort (N = 7,715; see Methods). Models were applied to 67,130 participants, and each compartment-specific predicted age gap (PAG) was defined as predicted minus chronological age after bias correction. We defined the Overall Aging Gap (OAG) as the unweighted mean of the eight PAGs. We then tested whether this composite captures a shared abdominal aging signal with broad prognostic value, whether disease-specific organ engagement patterns conditional on OAG provide biological refinement, and whether lifestyle contextualizes the resulting risk (Fig. 1).

**Figure 1.**
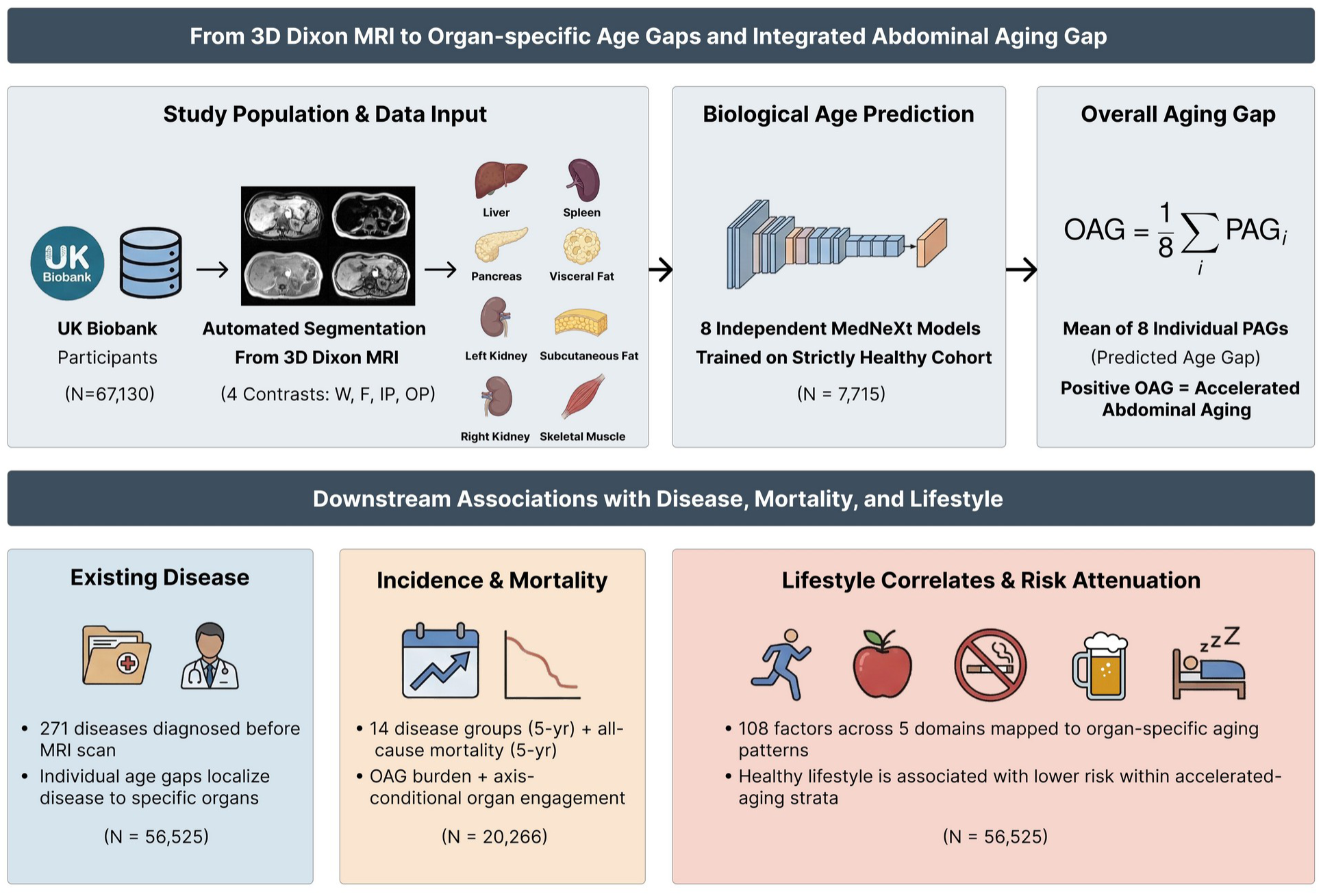
Study overview. Eight MedNeXt models trained on a strictly healthy subcohort (N = 7,715) estimated compartment-specific biological age from three-dimensional abdominal Dixon MRI in 67,130 UK Biobank participants. The Overall Aging Gap (OAG) was evaluated across four downstream domains: prevalent disease (N = 56,525); incident disease and all-cause mortality (N = 20,266; 5-year follow-up); and lifestyle factor associations (N up to 56,525) and HLS-stratified risk (endpoint-specific complete-HLS denominators).

### Abdominal age gaps are organized around a dominant shared axis

The ensemble of eight compartment models achieved a mean absolute error (MAE) of 2.69 years (R² = 0.816) in the held-out test set (N = 772). Per-compartment MAEs ranged from 2.75 years for liver to 3.81 years for visceral fat, with R² values of 0.60–0.78 across individual compartments (Fig. 2a, b), and the ensemble age estimate showed strong agreement with chronological age (Fig. 2c). Calibration slopes after bias correction were close to unity across all eight models (Supplementary Fig. S1).

**Figure 2.**
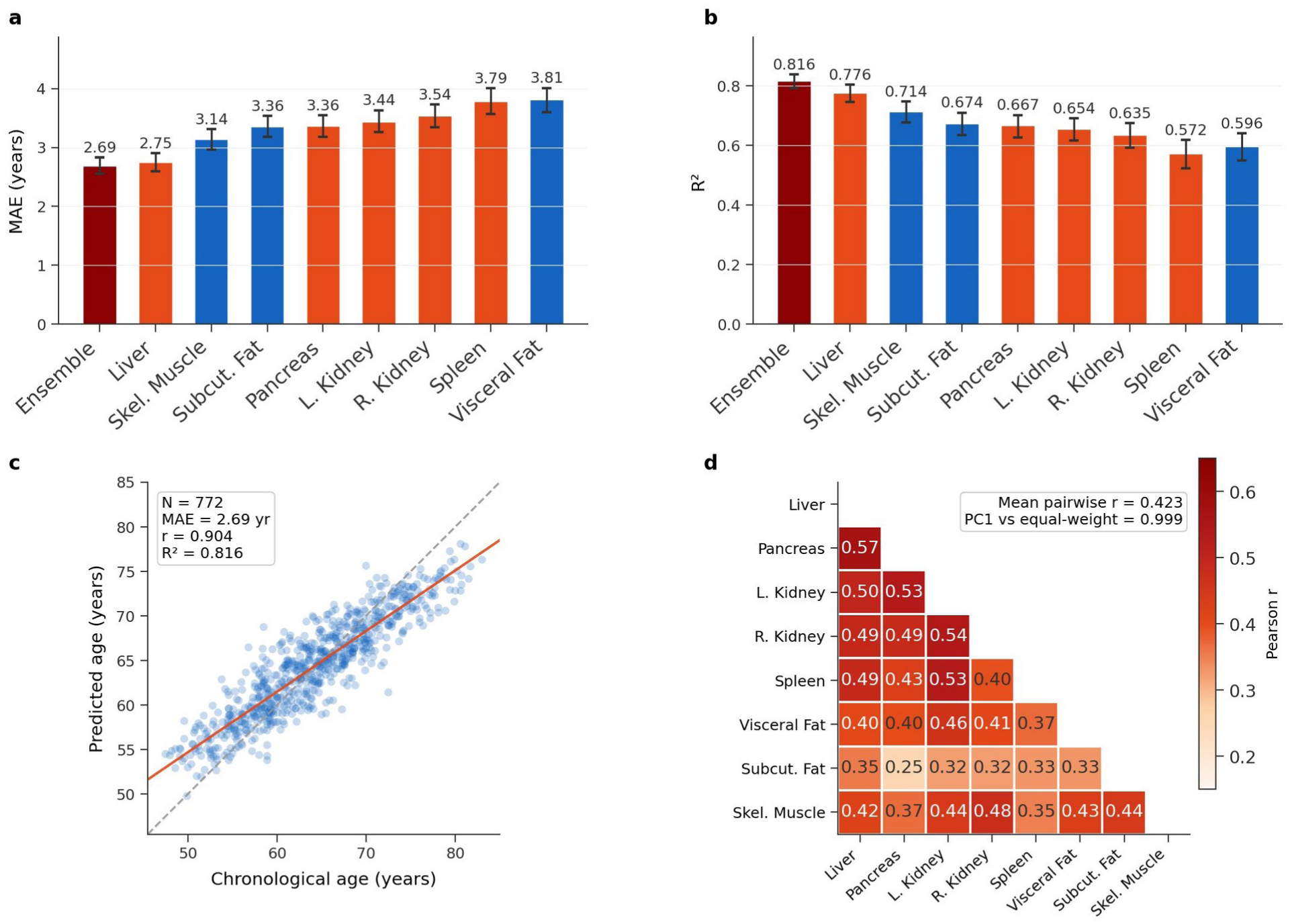
Model performance and derivation of the Overall Aging Gap. a, Per-compartment and ensemble mean absolute error (MAE) and b, R² in the held-out test set (N = 772). Error bars, 95% CI. c, Ensemble predicted versus chronological age. Dashed line, identity; red line, linear fit. d, Pairwise Pearson correlations among bias-corrected compartment-specific age gaps (mean r = 0.423; PC1-score correlation with the unweighted mean = 0.999).

All eight compartment-specific age gaps were positively intercorrelated (pairwise Pearson r = 0.253–0.569; mean r = 0.423; Fig. 2d), supporting their aggregation into a composite. The principal component spanned the eight compartments with approximately uniform loadings, and the unweighted mean of the eight PAGs (OAG) correlated almost perfectly with the first principal-component score (r = 0.999), supporting simple averaging as a faithful summary of the dominant shared axis. The first principal component accounted for approximately 50% of total age-gap variance (49.9% in the analytical cohort; 48.3% in the healthy subcohort), as expected from the observed mean pairwise correlation. The validation-set-derived bias correction was effective at the compartment level: in the covariate-complete subsample used for downstream analyses (N = 56,525), residual correlations between each compartment-specific PAG and chronological age were uniformly small (max |r| = 0.034 across the eight compartments), and partial correlations among the eight PAGs were essentially unchanged after additional adjustment for chronological age, sex and body-mass index (mean partial r = 0.408 controlling age and sex; mean partial r = 0.407 additionally controlling BMI; cf. raw mean r = 0.408 in the same subsample; Supplementary Fig. S2b). In the analytical cohort, OAG had a mean of 0.4 years (standard deviation = 2.5 years) and was only weakly correlated with chronological age (r = −0.031) and BMI (r = 0.082) after bias correction (Supplementary Fig. S2). In 4,789 participants with repeat imaging at a mean interval of 2.6 ± 1.0 years, OAG showed moderate test–retest concordance (ICC (3,1) = 0.706; Pearson r = 0.707; Supplementary Fig. S3), supporting the short-term stability of the composite measure.

### Prevalent analyses reveal broad remodeling with disease-specific abdominal patterns

We next examined how the hierarchical framework appears in established disease, interpreting prevalent associations as descriptive remodeling rather than formal prognostic refinement. As a population-level summary of cross-sectional disease burden, adjusted multimorbidity count rose monotonically across OAG deciles, from 3.57 distinct ICD-10 chapter A–N three-character categories in the lowest decile (D1) to 5.36 in the highest (D10) — a 1.50-fold gradient in adjusted disease burden — with a pronounced D9→D10 inflection that mirrored the non-linear OAG–mortality dose–response (Fig. 3a; P_trend = 6.3 × 10⁻¹⁵³; Poisson generalized linear model with robust standard errors, adjusted for age, sex and BMI). At the individual disease level, OAG was systematically elevated in prevalent disease, with significant associations for 271 of 430 ICD-10 chapter A–N three-character categories (FDR < 0.05 across diseases) spanning renal, respiratory, cardiovascular, metabolic, gastrointestinal, musculoskeletal, psychiatric and cancer categories. The median OAG difference among significant associations was 0.5 years, whereas eye and ear diseases, which lie outside the abdominal imaging field of view, were not significant. Among 16 representative aging-related diseases shown in Fig. 3b, the largest positive OAG differences were observed for end-stage renal disease (N19; +1.91 years), chronic obstructive pulmonary disease (J44; +1.85 years) and heart failure (I50; +1.62 years). Recurrent depression (F33; +1.61 years) also showed a notable positive association. Within this shared-axis background, individual diseases showed descriptive compartmental emphasis: chronic kidney disease showed predominant renal aging, chronic obstructive pulmonary disease showed broadly distributed acceleration across compartments, and type 2 diabetes showed greater acceleration in body-composition compartments than in solid organs (Fig. 3c; full compartment-level heatmap in Supplementary Fig. S4). Together, these cross-sectional results indicate that prevalent disease is associated with greater overall abdominal aging burden and disease-associated anatomical emphasis. Cross-modal biomarker phenotyping provided convergent support: higher OAG tertile aligned with adverse renal, glycemic and inflammatory profiles measured approximately 9 years before imaging (Supplementary Table S1; see Methods).

**Figure 3.**
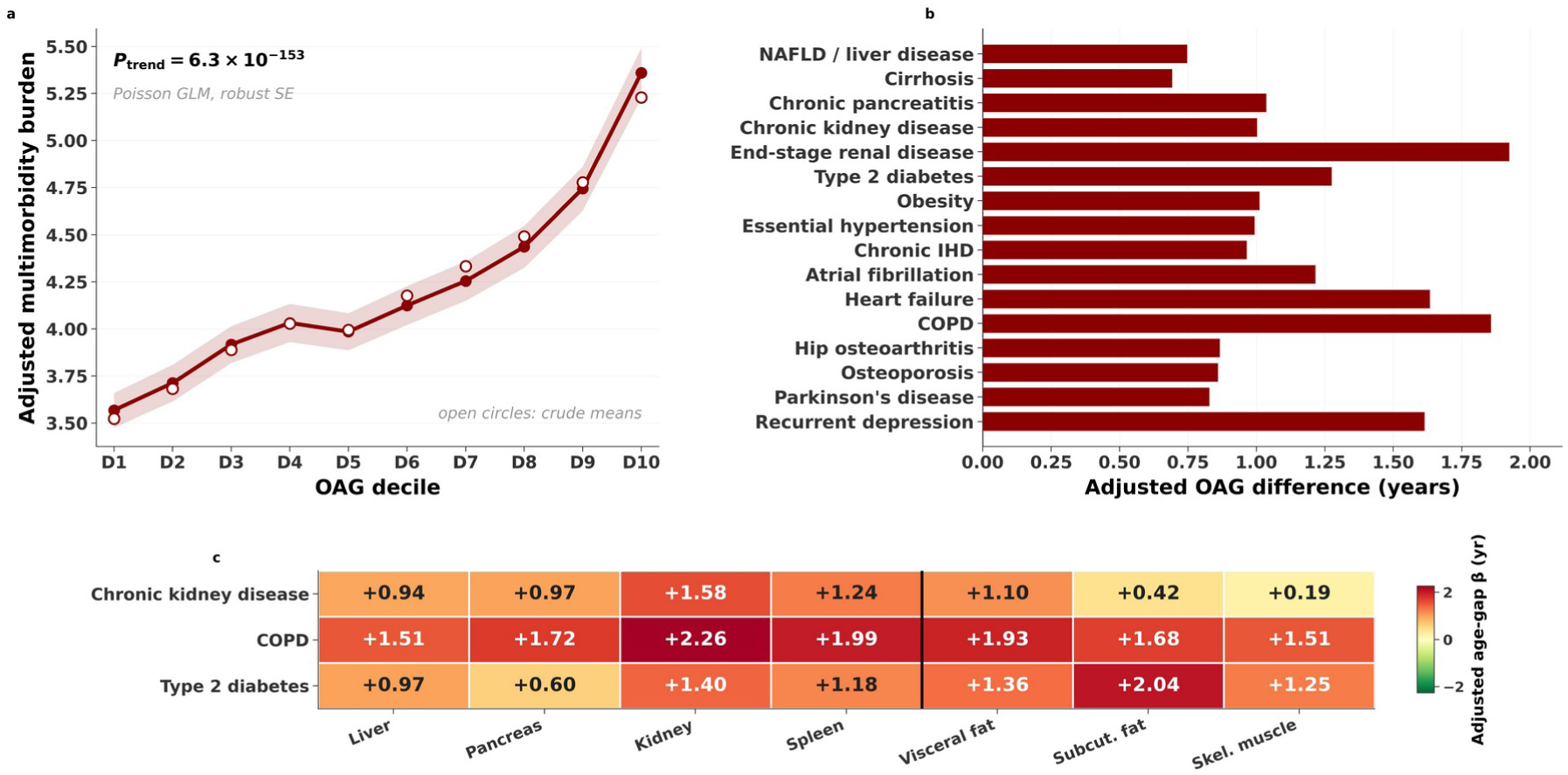
Prevalent disease associations with abdominal biological aging. a, Adjusted multimorbidity burden across OAG deciles (D1–D10) in the analytical cohort (N = 56,525); line and band, adjusted mean and 95% CI; open circles, crude per-decile means; P_trend tests the linear OAG-decile term. b, Adjusted OAG difference (years) for 16 representative aging-related diseases (OAG FDR < 0.05), ordered by anatomical system. c, Compartment-specific adjusted age-gap β (years) across seven functional compartments (left and right kidneys averaged into a single kidney PAG) for three archetypal diseases — chronic kidney disease (CKD), chronic obstructive pulmonary disease (COPD) and type 2 diabetes (T2D). All 271 diseases significant at FDR < 0.05 are shown in Supplementary Fig. S4.

### The shared axis stratifies broad prospective disease risk and all-cause mortality

In Cox proportional hazards models adjusted for age, sex, BMI, smoking and alcohol, OAG was associated with all 15 prespecified prospective endpoints — comprising 14 incident disease conditions and all-cause mortality — over follow-up of 5 years for disease incidence and 5 years for mortality (hazard ratios 1.15–1.49 per s.d.; FDR < 0.05 across endpoints; Fig. 4a). The strongest disease associations were observed for chronic obstructive pulmonary disease (HR = 1.49, 95% CI 1.34–1.66), atrial fibrillation (HR = 1.40), heart failure (HR = 1.38), type 2 diabetes (HR = 1.35) and abdominal cancer (HR = 1.30).

**Figure 4.**
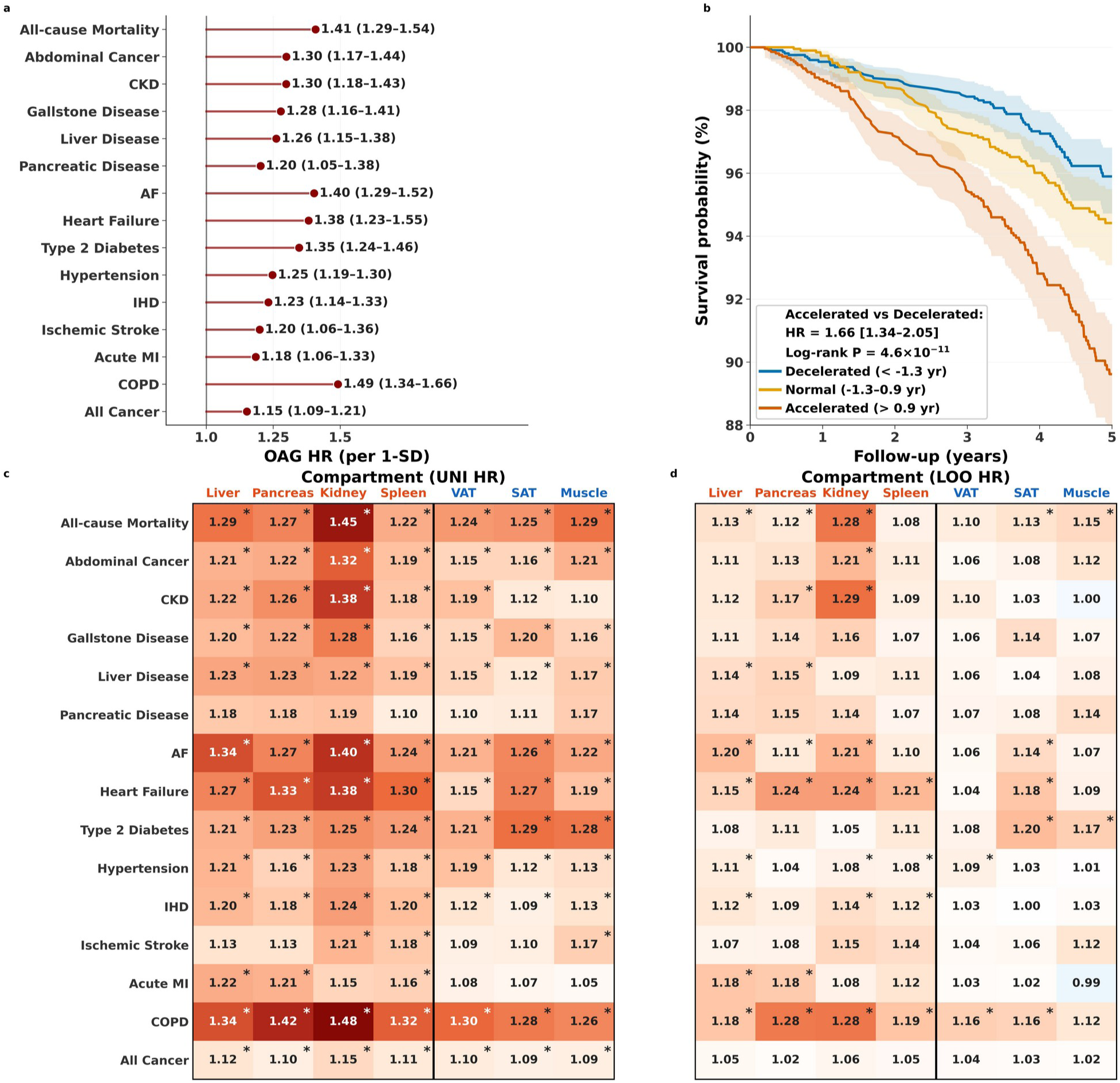
Prospective associations of OAG and axis-conditional organ engagement. a, Hazard ratios per s.d. of OAG for the 15 prospective endpoints (14 incident diseases and all-cause mortality, N = 20,266) over 5-year follow-up; adjusted for age, sex, BMI, smoking and alcohol. All endpoints reached FDR < 0.05. b, Kaplan–Meier survival curves for all-cause mortality by OAG tertile; shaded bands, 95% CI. Cox HR (accelerated vs decelerated) and log-rank P shown. c, Univariate compartment-level hazard ratios per s.d. of each functional compartment-specific PAG. d, Axis-conditional compartment-level hazard ratios from leave-one-out joint Cox models (LOO-OAG; see Methods). In c and d, the kidney column corresponds to the kidney PAG defined as the unweighted mean of bias-corrected left- and right-kidney PAGs, and heatmap rows follow the panel-a order. Stars, per-endpoint Benjamini–Hochberg FDR < 0.05 across the seven functional compartment tests. OAG–mortality dose–response spline: Extended Data Fig. 1.

Among these endpoints, all-cause mortality was followed for 5 years (1,250 deaths; HR = 1.41 per s.d., 95% CI 1.29–1.54; FDR < 0.001). Five-year survival was substantially lower in the accelerated OAG tertile compared with the decelerated tertile (Fig. 4b; Cox-adjusted HR (accelerated vs decelerated) = 1.66, 95% CI 1.34–2.05; log-rank P = 4.6 × 10⁻¹¹), corresponding to roughly twice the cumulative mortality at 5 years. Restricted cubic spline analyses indicated significant nonlinearity for all-cause mortality (P_nonlinearity = 8.3 × 10⁻⁶), with risk increasing more steeply at OAG values above approximately +4.6 years (Extended Data Fig. 1; full spline curves in Supplementary Fig. S5). In a separate broader-cohort mortality sensitivity analysis carried out in N = 40,985 participants drawn from the full analytical cohort (N = 56,525) using a narrower exclusion list restricted to serious prevalent disease at baseline (malignant neoplasms, ischemic heart disease, heart failure, cerebrovascular disease, chronic obstructive pulmonary disease, liver cirrhosis and chronic kidney disease), OAG remained associated with all-cause mortality (HR = 1.20, 95% CI 1.08–1.33; log-rank P = 3.5 × 10⁻⁸; Supplementary Fig. S6), confirming that the association is not driven by pre-existing high-mortality conditions.

### Axis-conditional organ engagement anatomically refines disease-specific vulnerability

To characterize whether disease-specific organ engagement provided refinement conditional on the shared axis, we fitted leave-one-out joint Cox models in which each functional compartment-specific PAG was evaluated alongside the leave-one-out OAG of the remaining six functional compartments (LOO-OAG). This design tests the compartment-specific contribution given the shared-axis signal captured by the other six functional compartments. By contrast, univariate compartment-level Cox models —each functional compartment-specific PAG entered alone with the same covariates—showed broadly significant compartment-by-endpoint associations across most endpoints (Fig. 4c), reflecting that each compartment individually indexes the shared aging axis and is therefore broadly prognostic on its own. After conditioning on LOO-OAG, the compartment-by-endpoint signal was substantially sparser, isolating disease-specific organ engagement beyond the shared axis (Fig. 4d). Per-endpoint Benjamini–Hochberg correction across the seven functional compartment tests identified at least one compartment with axis-conditional significance in 11 of 15 endpoints. The anatomical pattern of axis-conditional engagement varied across endpoints in biologically interpretable ways. Multi-system cardiorespiratory and metabolic conditions — chronic obstructive pulmonary disease, heart failure, atrial fibrillation, hypertension, ischemic heart disease, and all-cause mortality — engaged compartments spanning both solid organs (liver, pancreas, kidney, spleen) and body-composition tissues (visceral and subcutaneous adipose tissue, skeletal muscle), consistent with multi-system aging burden. Disease-specific solid-organ couplings reflected anatomically plausible patterns: chronic kidney disease engaged kidney and pancreas; acute myocardial infarction and liver disease each engaged liver and pancreas; and abdominal cancer engaged kidney. Type 2 diabetes engaged subcutaneous fat and muscle, consistent with adiposity- and sarcopenia-driven metabolic remodeling. The remaining endpoints — ischemic stroke, all cancer, pancreatic disease and gallstone disease — showed OAG association without compartment-specific refinement under the per-endpoint criterion.

These compartment-specific engagements were anatomically plausible and consistent with known disease biology. Several solid-organ engagements showed an additional contribution from the pancreas — consistent with pancreatic steatosis as a shared metabolic feature. Single-disease ICD-10 analyses further supported this anatomical structure: within the respiratory group, the detailed COPD code (J44) showed broad engagement across seven functional compartments, whereas emphysema (J43) showed more selective engagement restricted to three compartments. Full compartment-level statistics and ICD-10-level results are provided in Supplementary Tables S2 and S3.

### Lifestyle is associated with abdominal aging and with risk within accelerated-aging strata

As a translational extension, we next asked whether abdominal aging signals are associated with modifiable lifestyle factors and whether healthier lifestyle is associated with lower risk within accelerated-aging strata. Phenome-wide association analysis of 108 lifestyle factors across all eight compartment age gaps (Fig. 5a) showed that smoking-related factors — including pack-years, current smoking and passive smoke exposure — had the broadest associations with older-appearing compartments, whereas physical activity (vigorous, total, IPAQ group) had the broadest associations with younger-appearing compartments. Alcohol-related factors showed more compartment-specific patterns, with heavy spirits intake most strongly associated with liver and pancreas aging. Sleep duration showed a U-shaped relationship with OAG (quadratic P = 3.0 × 10⁻¹⁵; nadir ∼7.3 hours; Fig. 5b). All five lifestyle domains remained independently associated with OAG after mutual adjustment. Cross-sectionally, a five-domain Healthy Lifestyle Score (HLS) was associated with lower OAG (β = −0.187 per point, P < 10⁻⁹⁸; Cohen’s d = 0.27 across the HLS range).

**Figure 5.**
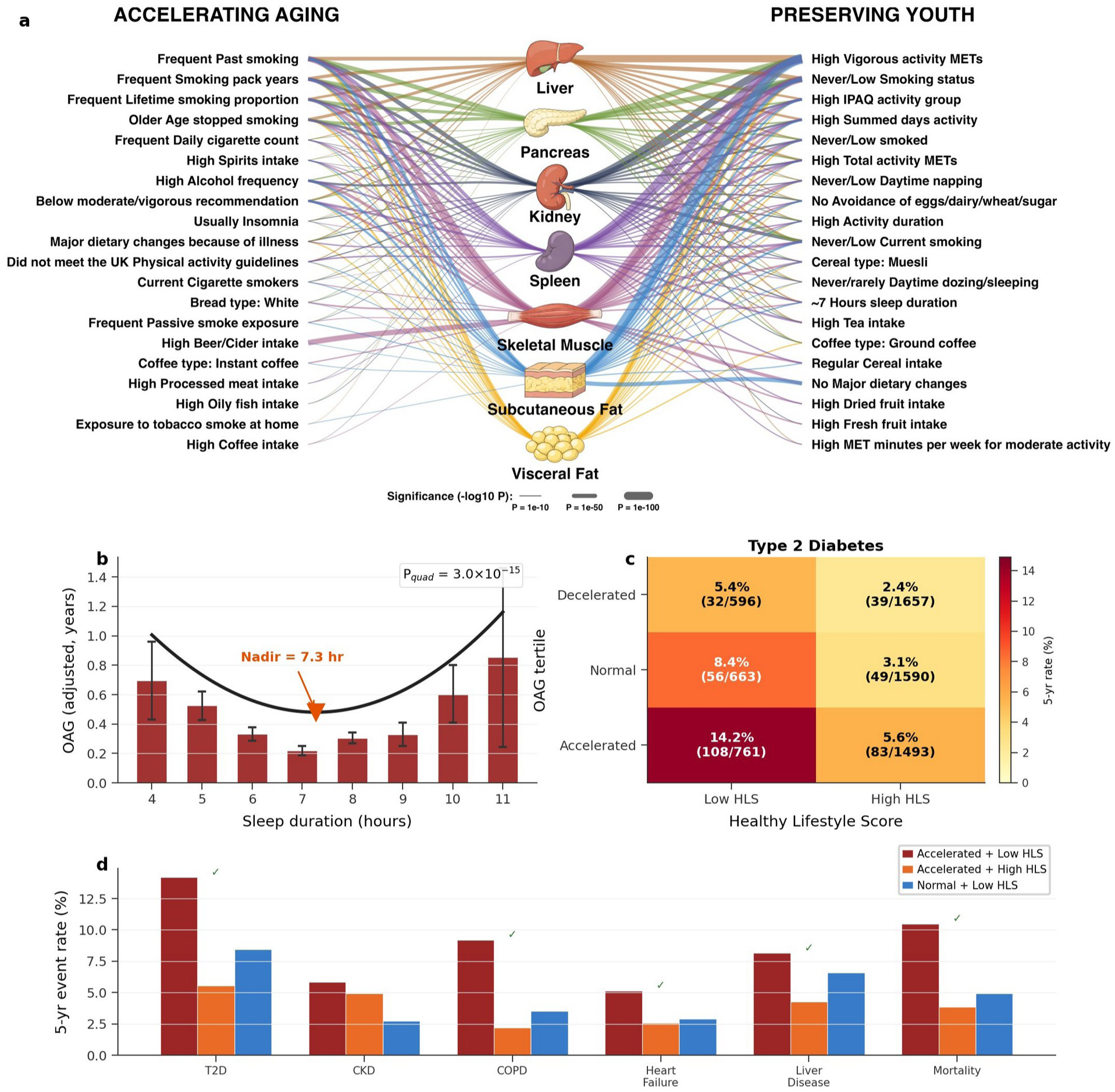
Lifestyle contextualizes abdominal aging burden and risk. a, Lifestyle–compartment association network. Edge thickness, −log₁₀(P); colors indicate the most strongly affected compartment. b, Adjusted relationship between sleep duration and OAG (P_quad = 3.0 × 10⁻¹⁵; nadir, 7.3 h). c, Five-year incidence of type 2 diabetes stratified by OAG tertile and Healthy Lifestyle Score (HLS) group (Low: 0–2; High: 3–5); cell values, incidence rate (events/N at risk). d, Five-year event rates across six representative endpoints by OAG × HLS strata. Dark red, T3 + Low; orange, T3 + High; blue, T2 + Low (reference). Green checks, lower-risk pattern (T3 + High at or below T2 + Low).

Among individuals with accelerated abdominal aging (highest OAG tertile), high HLS was associated with substantially lower observed 5-year incidence. For type 2 diabetes (Fig. 5c), 5-year incidence was 5.6% in the high-HLS subgroup versus 14.2% in the low-HLS subgroup — a 60% lower observed rate, lower than the 8.4% rate in the middle OAG tertile with low HLS. Across all 15 prospective endpoints (Extended Data Fig. 2; Supplementary Table S5), this lower-risk pattern was observed for 12 of 15 endpoints (80%), with a median relative difference of approximately 50% in the highest OAG tertile. Cox interaction tests reached nominal P < 0.05 for 6 of 15 endpoints and BH-FDR Q < 0.05 for COPD and all-cause mortality. The six representative endpoints shown in Fig. 5d span metabolic (T2D), renal (CKD), respiratory (COPD), cardiovascular (heart failure), hepatic (liver disease), and all-cause mortality domains. CKD was a notable exception (T3 + High HLS rate 4.9% > T2 + Low HLS rate 2.8%), consistent with the prominent renal-specific organ engagement signal in the axis-conditional refinement layer (Fig. 4d).

## Discussion

Within a single anatomically coherent abdominal MRI acquisition in 67,130 UK Biobank participants, we found that biological aging is hierarchically organized. Three observations define this hierarchy. First, the eight compartment-specific age gaps showed positive pairwise coupling (mean r = 0.42), with approximately half of total age-gap variance attributable to a dominant shared axis. Second, this shared axis, summarized as the unweighted compartment mean (Overall Aging Gap, OAG), broadly stratified prospective risk across all 15 endpoints (hazard ratios 1.15–1.49 per s.d.; mortality HR = 1.41 per s.d.). Third, after accounting for the shared axis, compartment-level associations became sparser and more anatomically coherent, identifying disease-specific organ engagement that refined endpoint risk at anatomically interpretable anchors without displacing the shared-axis signal. Structurally, these readouts are hierarchical: OAG summarizes the dominant shared axis, while compartment-level engagement is evaluated conditional on that axis. Functionally, they are complementary: OAG stratifies broad prospective risk, whereas axis-conditional compartment engagement provides disease-specific anatomical refinement. The substantial residual compartment-level variation that remains after accounting for the shared axis is what allows axis-conditional engagement to be tested in the first place, and clarifies why OAG and compartment engagement should not be treated as competing summaries of abdominal aging. Together, these findings establish a hierarchical framework for interpreting abdominal MRI age gaps rather than another abdominal composite score.

The hierarchical structure manifested in both established and incident disease, but with different statistical roles. In prevalent disease, OAG was systematically elevated and individual conditions showed compartment-level emphasis interpretable as disease-associated remodeling at the time of imaging. In incident disease, leave-one-out joint Cox models tested compartment-specific contributions conditional on the shared axis and identified axis-conditional engagement at anatomically interpretable anchors.

Our findings extend a growing literature on multi-organ biological aging in three specific ways. Prior multi-organ aging studies have introduced composite or organismal age measures alongside organ-specific clocks [1–4,15], including a recently developed methylation-based framework that summarizes 11 physiological systems [21]; however, these composites have typically been treated as parallel summaries rather than tested as a coordinated axis from which compartment-level deviations are conditionally evaluated. We extend this organizational logic to a single anatomically coherent imaging acquisition and operationalize it through formal axis-conditional decomposition. Second, recent multi-organ MRI aging clocks have estimated biological age across multiple body regions [3–6] without formally evaluating whether age gaps from a single anatomically coupled imaging acquisition share a dominant axis with compartment-level conditional refinement. Third, multi-organ aging frameworks have not systematically evaluated whether lifestyle further stratifies risk within accelerated-aging strata across diverse endpoints, an extension previously explored mainly in brain-age studies [7,16]. Our framework integrates these three contributions within a single imaging modality and study population.

The magnitude and structure of inter-compartment coupling we observe — mean pairwise r = 0.42, with approximately half of total age-gap variance attributable to a single dominant axis — provides quantitative grounding for this framework. This magnitude is closely consistent with the only directly comparable prior estimate in abdominal MRI: Le Goallec and colleagues reported a phenotypic correlation of 0.53 between liver- and pancreas-derived MRI age gaps in a two-dimensional study of two organs [8]. By contrast, plasma-proteomics-based aging clocks, which derive each organ score from a non-overlapping protein subset, report substantially weaker inter-organ coupling (mean r ≈ 0.16 across ten organ-enriched proteomic clocks in UK Biobank [18]; mean r ≈ 0.21 across eleven plasma proteomic organ clocks [2]). The intermediate-to-strong coupling we observe is therefore an expected feature of an integrated single-acquisition imaging setting that shares anatomical proximity, hemodynamic environment, and field of view, reflecting a coupled biological substrate — portal circulation, visceral-adipose-mediated inflammation, and organokine signaling — rather than a methodological artifact of overlapping imaging context. This biological interpretation justifies framing OAG as an integrated abdominal aging summary with axis-conditional compartment refinement, rather than as a substitute for organ-autonomous clocks.

Our findings have methodological implications for future multi-organ biological aging research. When multiple organ-age estimates are derived from the same individuals — whether from imaging, proteomic or methylation modalities — our results suggest that organ age gaps should not be reported solely as parallel summaries; the coupling structure among them, its magnitude and dimensional rank, and the disease-specific axis-conditional residuals, should itself be treated as a primary object of study. This perspective converges with a recent plasma-proteomics-based study of nine organ-specific aging signatures with 20-year follow-up in the Whitehall II cohort [17], in which individual age-related diseases were predicted by multiple organ aging signatures rather than mapping one-to-one onto a single organ — consistent with the broader principle that disease-specific multi-organ engagement, not organ-isolated aging, is a general feature of biological aging across measurement modalities. The kidney compartment is particularly informative in this context: in our axis-conditional analyses, kidney aging engaged the largest number of non-renal endpoints and showed the strongest lifestyle association of any compartment, suggesting that the kidney may serve as a multi-organ sentinel of cumulative metabolic, hemodynamic and inflammatory burden. Identifying compartments that integrate signal across multiple disease pathways — and characterizing the conditions under which they do — may be a productive direction for future organ-aging research aimed at biological substrates of broad prospective risk.

Beyond prognostic stratification, our lifestyle analyses address whether OAG can contextualize risk relative to modifiable factors. Healthier lifestyle was cross-sectionally associated with lower OAG, and within higher-aging strata it was associated with substantially lower subsequent disease risk in 12 of 15 endpoints (median ≈50% lower observed event rate within the highest OAG tertile). The longitudinal substudy clarified the temporal scope of these observations: the cross-sectional HLS–OAG association replicated at the second imaging visit (β = −0.132, P = 1.4 × 10⁻⁴), but within-person changes in HLS over the 2.6-year inter-scan interval were not associated with changes in OAG (β = −0.003, P = 0.93), largely because most participants did not substantively change lifestyle during this period. Within the same window, change in BMI was the only significant predictor of ΔOAG (β = −0.096, P = 7.5 × 10⁻⁷), indicating that abdominal aging trajectories did move with at least one quantifiable phenotypic change but at a temporal resolution that ΔHLS could not capture. Together, these observations support lifestyle as a contextualization layer for axis-level risk rather than evidence of intervention responsiveness, which awaits prospective and experimental tests. Kidney illustrates a complementary nuance of this layer: the kidney compartment was the most broadly responsive across non-renal disease and the most strongly associated with HLS, yet kidney disease itself showed the smallest lifestyle-associated risk offset once accelerated aging was established — consistent with the kidney’s known vulnerability to cumulative hemodynamic and metabolic injury [19,20], and demonstrating that composite and compartment-level readouts can provide complementary translational information within the same hierarchical framework.

Several limitations should be noted. The UK Biobank imaging cohort is affected by healthy volunteer bias, which limits generalizability, and all analyses were conducted within a single cohort, so independent external validation remains essential. OAG reflects abdominal rather than whole-body aging because the imaging field of view is anatomically restricted, and three of the eight compartments index body composition directly; although BMI was adjusted for, residual confounding cannot be excluded. We note, however, that partial correlations among the eight compartment-specific PAGs were essentially unchanged after additional adjustment for chronological age, sex and BMI (mean partial r = 0.407 vs raw mean r = 0.408 in the same covariate-complete subsample, a reduction of less than 0.5%), arguing against the OAG signal reducing to a body-habitus or chronological-age proxy. End-to-end models capture richer information than predefined imaging phenotypes but are less directly interpretable and will require dedicated explainability analyses. The stringent baseline exclusion strategy reduced the at-risk cohort for incident analyses to approximately 20,000 participants; per-endpoint exclusion may provide complementary estimates. Lifestyle stratification analyses were not adjusted for socioeconomic factors such as Townsend deprivation index or education, so observed associations may partly reflect social advantage rather than direct biological effects. Repeat imaging supported moderate short-term stability of OAG, but longer-term within-person trajectories remain unknown; the ongoing expansion of repeat abdominal MRI in the UK Biobank may provide the statistical power and temporal leverage needed to test whether sustained lifestyle change prospectively alters abdominal aging trajectories. OAG was defined as a simple unweighted mean of eight compartments to prioritize interpretability and robustness; in this construction bilateral kidneys contribute 2/8 = 25% of the OAG weight, and alternative weighting strategies (e.g., variance-weighted or PCA-derived composites) merit future comparison. Although renal ICD codes were excluded in the kidney sensitivity analysis, kidney age gap may still partly reflect subclinical renal dysfunction; future work should test whether its associations persist after adjustment for baseline eGFR. With external validation and longer-term longitudinal confirmation, this hierarchical framework — shared abdominal aging burden plus axis-conditional compartment engagement — could move abdominal imaging from a parallel set of organ clocks toward an integrated readout that combines broad prospective risk stratification with anatomically resolved disease-specific signals.

## Methods

### Study population

We analyzed abdominal magnetic resonance imaging (MRI) data from the UK Biobank, a prospective population-based cohort of approximately 500,000 participants recruited at ages 40–69 years between 2006 and 2010. A subset underwent abdominal MRI as part of the UK Biobank Imaging Study. The primary analyses used the baseline imaging assessment (instance 2); a separate repeat-imaging cohort (instance 3) was used exclusively for test–retest reliability analyses (see Longitudinal stability below). Of 69,546 participants with abdominal MRI at the baseline imaging assessment, 67,130 had successful automated segmentation and age predictions for all eight target compartments. Participants with incomplete imaging data, motion artifacts, or failed segmentation for any compartment were excluded. Requiring non-missing BMI, smoking and alcohol status yielded 64,240 participants. Model development/evaluation participants — comprising training, validation and held-out test sets (combined N = 7,715) — were then excluded to prevent leakage between model development and downstream analyses. The primary analytical cohort for prevalent disease, biomarker and lifestyle analyses comprised N = 56,525. The prospective endpoint cohort comprised N = 20,266 after additional blanket prevalent-disease exclusion (see Incident disease and mortality analyses below) and was used for both incident disease and all-cause mortality analyses, supporting a single consistent at-risk design across all 15 prospective endpoints. A separate broader-cohort mortality sensitivity analysis was carried out in N = 40,985 participants drawn from the analytical cohort using a narrower exclusion list (see Incident disease and mortality analyses). A study flow diagram is provided in Supplementary Fig. S11.

### Abdominal MRI acquisition

Abdominal images were acquired on 1.5 T Siemens MAGNETOM Aera scanners at dedicated UK Biobank imaging centers using the standard neck-to-knee Dixon VIBE protocol. This acquisition yields four contrast volumes from a single scan: in-phase (IP), opposed-phase (OP), water-only (W), and fat-only (F). All four channels were used as multi-channel inputs to the deep learning models, providing complementary information on organ parenchyma, lipid distribution, and tissue boundaries.

### Organ and tissue segmentation

Eight abdominal compartments were segmented from the Dixon MRI volumes: liver, pancreas, left kidney, right kidney, spleen, visceral adipose tissue (VAT), subcutaneous adipose tissue (SAT), and skeletal muscle. Automated segmentation was performed using TotalVibeSegmentator [13], a deep learning framework for high-throughput Dixon MRI segmentation, building on the nnU-Net-based abdominal organ segmentation pipeline [14]. Segmentation quality was assessed using an automated quality-control pipeline; participants were excluded if any target structure yielded an empty mask.

### Biological age prediction models

For each of the eight compartments, we trained an independent biological age prediction model based on a modified MedNeXt architecture [12] with multi-stream input fusion. Each Dixon contrast channel was processed through a stem block (3 × 3 × 3 convolution, Group Normalization, GELU), followed by cross-attention fusion (embedding dimension 32; 4 attention heads) and four MedNeXt stages (channel widths 32, 64, 128, 256) with 7 × 7 × 7 depth-wise separable convolutions. Spatial-and-channel squeeze-and-excitation (scSE) modules and Dynamic Affine Feature Transform (DAFT) sex conditioning were incorporated. For solid organs (liver, pancreas, kidneys, spleen), input volumes were cropped to the organ bounding box with a 10-voxel margin. For body-composition compartments (VAT, SAT, skeletal muscle), the axial field of view was standardized to the region between the superior extent of the liver and the inferior extent of the coccyx, determined automatically from the segmentation masks. All volumes were resampled to 2.2 × 2.2 × 3.0 mm and center-cropped or padded to 96 × 96 × 96 voxels. Models were trained using mean absolute error loss with the AdamW optimizer for 300 epochs. Full training details, including data augmentation, batch size, learning rate schedule and early stopping criteria, are provided in Supplementary Methods.

### Healthy cohort definition

The training cohort (N = 7,715; train/val/test = 6,172/771/772) was defined by excluding participants with any condition known to affect abdominal organ morphology. Exclusion criteria combined linked hospital inpatient records, self-reported health data, and imaging-derived phenotypes: (1) any hospital inpatient ICD-10 diagnosis (category 41270) recorded up to and including the imaging visit; (2) self-reported long-standing illness, disability, or infirmity (field 2188); (3) self-reported diabetes (field 2443); (4) self-reported stroke history (field 4056); (5) self-reported health rated less than “good” (field 2178); and (6) liver proton density fat fraction (PDFF) ≥ 5.5% or pancreas PDFF ≥ 6.2%, to exclude subclinical hepatic steatosis and pancreatic fat infiltration. A broader reference cohort (N = 9,373), defined using less stringent criteria (excluding only type 2 diabetes and active abdominal malignancy), was used to provide the standard deviation for downstream z-score normalization of OAG and compartment-specific PAGs; bias-correction parameters themselves were estimated exclusively in the validation subset (see “Bias correction, terminology, and OAG definition” below). This larger reference was chosen to provide more stable variance estimates for standardization, while the stricter training cohort ensured that age-prediction models were not influenced by disease-related morphological changes. Train, validation, and test splits were performed at the participant level to prevent data leakage across sets. The held-out test set comprised 772 participants used for both per-compartment and ensemble-level evaluation.

### Bias correction, terminology, and OAG definition

For each compartment, the model outputs a raw predicted age. The raw age gap is defined as raw predicted age minus chronological age. A linear bias correction was fitted on the validation set only, modeling mean raw age gap as a function of chronological age; correction parameters were estimated exclusively from validation participants and then frozen before application to the held-out test set and all downstream analyses. The bias-corrected predicted age gap (PAG) is the residual after this correction. This procedure removes regression-to-the-mean bias while preserving inter-individual variation attributable to biological or clinical factors and ensures no information from the test or analytical cohorts influenced the correction.

The Overall Aging Gap (OAG) was defined as the unweighted mean of the eight compartment-specific bias-corrected PAGs, motivated by consistently positive intercorrelations (mean pairwise r = 0.423), approximately uniform PC1 loadings, and the near-perfect correlation between OAG and the first principal-component score (r = 0.999). The eight compartments comprised liver, pancreas, left kidney, right kidney, spleen, visceral adipose tissue, subcutaneous adipose tissue and skeletal muscle; bilateral kidneys therefore contributed two of the eight components to OAG (effective weight 2/8 = 25%), reflecting their treatment as anatomically distinct prediction targets in the segmentation and modeling pipeline. For axis-conditional compartment-engagement analyses (see “Leave-one-out joint Cox models” below), for the kidney cross-disease sensitivity analysis, and for visualization of compartment-level disease patterns in Fig. 3c, Fig. 5a and Supplementary Figs. S4 and S8, we additionally defined a kidney PAG as the unweighted mean of the bias-corrected left- and right-kidney PAGs, yielding seven functional compartments (liver, pancreas, kidney, spleen, visceral adipose tissue, subcutaneous adipose tissue and skeletal muscle) and seven functional compartment tests per endpoint. This kidney-merging step applies only to the analyses listed above; OAG itself is the eight-compartment mean throughout. Alternative weighting strategies (e.g., variance-weighted or PCA-derived composites) merit future comparison.

The ensemble prediction, reported in model performance analyses (Fig. 2a, b), refers to the mean of the eight raw predicted ages (before bias correction) and is distinct from OAG. All downstream disease, biomarker, lifestyle and mortality analyses used OAG (bias-corrected).

### Longitudinal stability

To assess test–retest reliability, we identified 4,789 participants who completed both the baseline imaging assessment (instance 2) and a repeat imaging assessment (instance 3; mean interval 2.6 ± 1.0 years). The same frozen bias-correction parameters derived from the baseline validation set were applied to repeat-imaging predictions before computing PAGs and OAG. Concordance between baseline and repeat OAG was assessed using Pearson correlation and intraclass correlation coefficient (ICC (3,1), two-way mixed model, consistency definition). The inter-scan interval was calculated as the difference between scan ages at the two visits.

### Prevalent disease associations

Cross-sectional associations between OAG and 688 ICD-10-coded prevalent diseases were initially screened using ordinary least squares regression with OAG as the dependent variable and disease status as the predictor, adjusted for age, sex, and BMI. The primary disease-association test family was restricted to the 430 ICD-10 chapter A–N three-character categories meeting the prevalent-disease ascertainment criteria (after the chapter filter that removes ICD-10 chapters covering external causes, codes for health-status factors, and other non-disease classifications); Benjamini–Hochberg FDR correction was applied across these 430 tests, of which 271 reached FDR < 0.05. Organ-level associations were similarly assessed for each compartment-specific PAG. To summarize cross-sectional disease burden as a function of OAG, we additionally fitted an adjusted multimorbidity-gradient model (Fig. 3a). For each participant, multimorbidity was defined as the count of distinct ICD-10 chapter A–N three-character categories meeting the prevalent-disease ascertainment criteria (430 codes retained after the chapter filter). OAG was binned into deciles in the analytical cohort (N = 56,525). Decile-specific adjusted means and 95% confidence intervals were estimated by g-computation from a Poisson generalized linear model fitted as multimorbidity_count ∼ C(OAG_decile) + age + sex + BMI with HC3 robust standard errors and delta-method variance propagation across the decile contrasts. A parallel Poisson model with OAG decile entered as a continuous term provided a linear-trend test (P_trend). The model was overdispersed (variance/mean ratio = 3.69), as expected for multimorbidity counts; HC3 robust standard errors accommodate this overdispersion. Sensitivity analyses (excluding I10 essential hypertension; using the raw 22-disease curated count; negative-binomial regression) are reported in Supplementary Methods.

### Incident disease and mortality analyses

Incident disease analyses used Cox proportional hazards models with OAG (z-scored by the reference-cohort standard deviation) as the primary predictor, adjusted for centered age, sex, z-scored BMI, smoking status, and alcohol status (N = 20,266; 5-year follow-up). To approximate a disease-free baseline, participants diagnosed before the imaging visit with serious acute conditions (malignant neoplasms, ischemic heart disease, myocardial infarction, heart failure, stroke, COPD, severe asthma), common chronic diseases (hypertension, atherosclerosis, aneurysm, dyslipidemia, obesity, hypothyroidism, anemias, rheumatoid arthritis) or abdominal/digestive diseases were excluded; training-set participants were also excluded. This blanket prevalent-disease exclusion, rather than per-endpoint removal, was chosen to approximate a homogeneous sub-healthy baseline, thereby reducing the influence of pre-existing multimorbidity on incident risk estimates while retaining a standard Cox at-risk design in which all remaining participants contributed person-time until the event of interest or censoring. Fifteen prospective endpoints were defined, comprising 14 incident disease conditions ascertained from linked hospital inpatient ICD-10 records — spanning cardiovascular, metabolic, renal, hepatic, pancreatic, respiratory, gastrointestinal and oncological categories — and all-cause mortality. Endpoints with weak mechanistic linkage to abdominal aging on the basis of anatomy or prior literature (dementia, cataract/glaucoma, hearing loss, musculoskeletal disorders, diverticular disease, depression and gastroesophageal reflux disease) were excluded a priori from the primary endpoint family to focus the analysis on conditions with anatomically plausible coupling to abdominal compartment biology. Full ICD-10 code lists are provided in Supplementary Table S4. A 180-day exclusion window after imaging was applied to reduce reverse-causation bias. Hazard ratios are reported per standard deviation of OAG. The proportional hazards assumption was assessed using Schoenfeld residuals; no substantial violations were detected.

All-cause mortality was analyzed using up to 5 years of follow-up with the same covariate adjustment as the incident disease models, in the same prospective endpoint cohort (N = 20,266) used for incident disease analyses. Follow-up time was measured from the imaging visit to death (from linked national death registries) or administrative censoring, whichever came first. Kaplan–Meier curves were stratified by OAG tertiles with log-rank tests. Restricted cubic splines (4 knots at the 5th, 35th, 65th, and 95th percentiles) were used to assess nonlinearity; the inflection point was defined as the OAG value at which the second derivative of the log-hazard ratio was maximized, restricted to the central 90% of the data range. As a separate broader-cohort mortality sensitivity analysis, we repeated the mortality model in N = 40,985 participants drawn from the full analytical cohort (N = 56,525) using a narrower exclusion list — only serious prevalent disease at baseline (malignant neoplasms, ischemic heart disease, heart failure, cerebrovascular disease, chronic obstructive pulmonary disease, liver cirrhosis and chronic kidney disease) — rather than the broader prospective-endpoint exclusion list applied to the N = 20,266 cohort. This sensitivity analysis is therefore performed in a different, larger cohort than the primary mortality analysis and is reported separately.

### Leave-one-out joint Cox models for axis-conditional compartment engagement

To characterize axis-conditional organ engagement, we fitted leave-one-out joint Cox models in which each functional compartment-specific PAG was entered alongside the leave-one-out OAG formed from the remaining six functional compartments (LOO-OAG_−c_), as z-scored predictors, alongside the same covariates used in the primary incidence analyses (centered age, sex, z-scored BMI, smoking and alcohol status). The partial Wald P-value for PAG_c_ provides an axis-conditional test of organ engagement: the compartment-specific hazard ratio given the shared-axis signal captured by the other six functional compartments.

Formally, for each participant *i* and compartment *c* we defined the leave-one-out OAG as the unweighted mean of the six remaining functional compartment-specific PAGs,

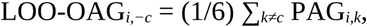

and the axis-conditional Cox model for endpoint *d* was specified as

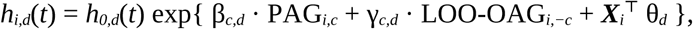

where β*_c,d_* is the axis-conditional log-hazard ratio for compartment *c* and endpoint *d* (the primary parameter of interest reported in Fig. 4d), γ*_c,d_* captures the residual shared-axis signal carried by the other six functional compartments, and ***X****_i_* denotes the covariate vector (centered age, sex, z-scored BMI, smoking and alcohol status). The partial Wald test on β*_c,d_* provides the axis-conditional test of organ engagement reported in Fig. 4d, while jointly estimating β*_c,d_* and γ*_c,d_* avoids the part-whole circularity that would arise from conditioning PAG*_c_* on OAG (which itself contains PAG*_c_*).

Because the kidney PAG was defined as the unweighted mean of left- and right-kidney bias-corrected PAGs from the start (see “Bias correction, terminology, and OAG definition”), the kidney enters axis-conditional analyses as a single functional compartment with unit weight. We applied per-endpoint Benjamini–Hochberg FDR correction across the seven functional compartment tests as the primary test family for axis-conditional compartment engagement (most relevant for a disease-specific organ-engagement question, and more powerful than global correction across all endpoints); global FDR correction across all endpoints × compartments was retained as a sensitivity analysis. Note that this per-endpoint correction across compartments is distinct from the FDR correction applied to OAG associations across endpoints (Statistics and reproducibility). Compartment-specific engagement is reported descriptively at the per-endpoint level, listing which compartments reached axis-conditional significance for each endpoint; for grouping purposes in the Results, we informally distinguish solid-organ compartments (liver, pancreas, kidney, spleen) from body-composition compartments (visceral adipose tissue, subcutaneous adipose tissue, skeletal muscle), reflecting anatomical and physiological structure rather than a formal classification scheme.

### Kidney cross-disease sensitivity analysis

To assess whether the kidney compartment showed broad cross-disease associations independent of renal-specific endpoints, we ranked the seven functional compartments by adjusted effect size across non-renal diseases. The kidney PAG (defined as the unweighted mean of bias-corrected left- and right-kidney PAGs; see “Bias correction, terminology, and OAG definition”) was used as the primary kidney measure. As a secondary check, left and right kidney PAGs were also evaluated separately, and we report the kidney with the larger absolute effect size for each disease (denoted “best of left and right” where this comparison is shown). Kidney-specific ICD-10 categories (N17–N28, I12–I13, C64–C68) were excluded to avoid tautological associations. Rankings were computed separately for prevalent associations and prospective incidence analyses.

### Biomarker and medication phenotyping

Thirty-two blood biomarkers from the UK Biobank baseline assessment (collected approximately 9–10 years before the imaging visit) were compared across OAG tertiles. To isolate the independent association between OAG and each biomarker, both OAG and the biomarker were residualized on age, sex, and BMI before computing partial Spearman correlations and adjusted Cohen’s d between the highest and lowest residualized-OAG tertiles. Tertile labels (T1, T2, T3) in figures refer to these residualized-OAG tertiles. Benjamini–Hochberg FDR correction was applied across all 32 tests.

### Healthy Lifestyle Score construction

A Healthy Lifestyle Score (HLS, range 0–5) was constructed from five lifestyle domains assessed at the imaging visit (UK Biobank instance 2): physical activity, smoking status, alcohol consumption, diet, and sleep duration, following established 5-domain healthy-lifestyle frameworks in UK Biobank [22]. Each domain contributed one point if the participant met a low-risk criterion (0 otherwise), giving an integer score from 0 (least healthy) to 5 (most healthy). Low-risk criteria were: (i) physical activity — meeting the World Health Organization recommendation of ≥150 min/week of moderate or ≥75 min/week of vigorous activity (or equivalent combination), based on the International Physical Activity Questionnaire (IPAQ); (ii) smoking — never or former smoker; (iii) alcohol — ≤14 UK units/week, consistent with current UK Chief Medical Officer guidelines; (iv) diet — a healthy-diet score of ≥4 of 7 items (≥3 servings of fruit/day, ≥3 servings of vegetables/day, ≥2 servings of fish/week, ≤2 servings of processed meat/week, ≤1 serving of unprocessed red meat/day, refined-grain intake below cohort median, whole-grain intake above cohort median); and (v) sleep duration — self-reported habitual sleep of 7–8 hours per night. Participants were grouped into Low HLS (0–2 points) and High HLS (3–5 points) for lifestyle-stratified risk analyses (Fig. 5c, d). Participants with missing data on any single lifestyle domain were excluded from HLS-based analyses. The complete-HLS cohort comprised N = 56,525 for cross-sectional lifestyle analyses and N = 20,266 for prospective lifestyle-stratified risk analyses; endpoint-specific denominators are shown in Supplementary Table S5.

### Lifestyle × OAG interaction

To assess whether healthier lifestyle was associated with lower disease risk among individuals with accelerated aging, participants were cross-classified by OAG tertile (T1, T2, T3) and HLS group (Low: 0–2; High: 3–5). Five-year incidence rates were computed for each of the six resulting cells across all 15 prospective endpoints (5-year follow-up for all-cause mortality). The key comparison tested whether the highest OAG tertile with high HLS (T3 + High) showed disease rates at or below the middle tertile with low HLS (T2 + Low), indicating an apparent lower-risk pattern roughly equivalent to one tertile of biological aging burden. Formal interaction was tested by adding an OAG × HLS product term to Cox proportional hazards models adjusted for age, sex, and BMI. Smoking and alcohol were not included as additional covariates in the interaction models because both are components of the HLS composite, and their inclusion would constitute partial conditioning on the exposure. To identify which lifestyle domains contributed most in individuals with accelerated aging, domain-specific regression coefficients were compared across OAG tertiles.

### Ethics

This research was conducted using the UK Biobank Resource under Application Number 132578. UK Biobank received ethical approval from the North West Multi-Centre Research Ethics Committee (Ref: 11/NW/0382). All participants provided written informed consent at enrollment. The study was performed in accordance with the Declaration of Helsinki.

### Statistics and reproducibility

No formal sample-size calculation was performed; the analytical cohort was determined by UK Biobank imaging availability and data completeness. No data were excluded beyond predefined imaging quality control, cohort definition, and analysis-specific covariate or endpoint eligibility criteria. All statistical tests were two-sided. Multiple testing was controlled using Benjamini–Hochberg false discovery rate (FDR) correction applied separately within each analysis domain. For OAG associations (prevalent disease, incident disease, biomarker, medication and lifestyle associations), FDR correction was applied across endpoints within each domain. For axis-conditional organ engagement, per-endpoint FDR correction across the seven functional compartment tests was applied as the primary test family, with global FDR correction across all endpoints × compartments retained as a sensitivity analysis. Effect sizes are reported with 95% confidence intervals throughout. The study is observational; investigators were not blinded to participant characteristics during data analysis, as blinding is not applicable to secondary analyses of a population cohort. Deep learning model training, bias correction, and statistical analysis pipelines were defined before the final outcome analyses were run. All analyses were conducted in Python 3.10; code is available from the corresponding authors upon reasonable request after publication.

### Statistical software

Deep learning models were implemented using the PyTorch framework with the MONAI library for medical imaging. Statistical analyses were performed in Python 3.10 using lifelines v0.27, statsmodels v0.14 and scipy v1.11. Visualizations were generated with matplotlib v3.8.

## Declarations

Supplementary information Supplementary figure legends are provided at the end of this manuscript; supplementary figure images and tables accompany the manuscript online.

Acknowledgements This research was conducted using the UK Biobank Resource under Application Number 132578. We thank all UK Biobank participants and staff.

Funding No specific funding was received for this work.

Competing interests The authors declare no competing interests.

Ethics approval UK Biobank received ethics approval from the North West Multi-Centre Research Ethics Committee (Ref: 11/NW/0382). All participants provided written informed consent. Full ethics statement is provided in the Ethics subsection of Methods.

Data availability Individual-level imaging and health data were accessed through the UK Biobank Resource (Application Number 132578) and are available to approved researchers via https://www.ukbiobank.ac.uk. The derived Overall Aging Gap and compartment-specific predicted age differences used in the statistical analyses will be returned to UK Biobank for use by other researchers upon publication.

Code availability Code is available from the corresponding authors upon reasonable request after publication.

## Extended Data Figure Legends

**Extended Data Fig. 1 | OAG–mortality dose–response.** Restricted cubic spline showing the hazard ratio for all-cause mortality as a function of continuous OAG (4 knots at the 5th, 35th, 65th and 95th percentiles), with 95% confidence band; P_nonlinearity = 8.3 × 10⁻⁶; the inflection point at approximately +4.6 years marks where the second derivative of the log-hazard is maximized within the central 90% of the data range. Referenced in Results: shared axis predicts incident disease and all-cause mortality; Fig. 4 caption.

**Extended Data Fig. 2 | Lifestyle-stratified 5-year incidence across all 15 prospective endpoints.** Five-year event rates by OAG tertile (T1, T2, T3) × HLS group (Low: 0–2; High: 3–5) for each of the 14 incident disease endpoints and all-cause mortality, complementing the six representative endpoints shown in Fig. 5d. Cells highlight the lower-risk pattern in T3 + High HLS relative to T2 + Low HLS where present. Referenced in Results: lifestyle is associated with abdominal aging and with risk within accelerated-aging strata.

## Supplementary Figure Legends

**Supplementary Fig. S1 | Calibration slopes after bias correction.** Per-compartment calibration slopes (predicted vs. chronological age) after applying the validation-set-derived linear bias correction. Slopes should be close to unity across all eight models. Referenced in Results: Model performance.

**Supplementary Fig. S2 | OAG correlation with chronological age and BMI after bias correction.** Scatter plots showing weak residual correlations between bias-corrected OAG and chronological age (r = −0.031) and BMI (r = 0.082) in the analytical cohort. Referenced in Results: Model performance.

**Supplementary Fig. S2b | Inter-compartment PAG correlation matrices: raw, partial controlling age and sex, and partial controlling age, sex and BMI.** Three-panel 8 × 8 heatmap of pairwise Pearson correlations among the eight bias-corrected compartment-specific age gaps in the covariate-complete analytical subsample (N = 56,525), under three specifications: raw Pearson on bias-corrected PAG (matched to Le Goallec 2022 [8], Oh 2023 [2] and Wang/Xiao 2026 [18]); partial Pearson controlling chronological age and sex (matched to Tian 2023 [1]); and partial Pearson additionally controlling BMI. Mean off-diagonal r is annotated on each panel. The three matrices are nearly indistinguishable (mean r = 0.408, 0.408 and 0.407 respectively), demonstrating that inter-compartment coupling is not driven by residual chronological-age dependence, sex or BMI. A companion table reports per-compartment residual correlations of each PAG with chronological age, sex and BMI; the maximum |r| with chronological age across the eight compartments was 0.034. Referenced in Results: shared axis is organized around a dominant shared axis; Discussion: cross-organ coupling.

**Supplementary Fig. S3 | Test–retest concordance of OAG and compartment-specific age gaps.** Scatter plot of OAG at Visit 2 vs. Visit 3 (N = 4,789; ICC(3,1) = 0.706; Pearson r = 0.707; mean interval 2.6 ± 1.0 years). Per-compartment ICCs may also be shown. Referenced in Results: Model performance.

**Supplementary Fig. S4 | Full compartment-level prevalent disease heatmap.** Heatmap of mean age-gap differences (years) across seven functional compartments, with left and right kidneys averaged into a single kidney PAG, for all 271 significant prevalent diseases (FDR < 0.05). Extends the representative subset shown in Fig. 3. Referenced in Results: Prevalent disease.

**Supplementary Fig. S5 | Restricted cubic spline dose–response curves.** Full restricted cubic spline curves showing the hazard ratio for all-cause mortality (and optionally key incident disease endpoints) as a function of continuous OAG, with 95% confidence bands. Extends Extended Data Fig. 1. Referenced in Results: shared axis predicts incident disease and all-cause mortality.

**Supplementary Fig. S6 | Healthy-baseline mortality sensitivity analysis.** Kaplan–Meier cumulative mortality by OAG tertile in the separate broader-cohort mortality sensitivity analysis (N = 40,985 participants drawn from the full analytical cohort N = 56,525, using a narrower exclusion list restricted to serious prevalent disease at baseline; log-rank P = 3.5 × 10⁻⁸; adjusted HR (T3 vs T1) = 1.20 [1.08–1.33]). Referenced in Results: shared axis predicts incident disease and all-cause mortality.

**Supplementary Fig. S7 | Full per-disease organ engagement heatmap.** Compartment-level hazard ratios (rows = 15 endpoints; columns = 7 functional compartments with kidneys merged) from leave-one-out joint Cox models, with per-endpoint FDR significance indicators. Supports Fig. 4d. Referenced in Results: axis-conditional organ engagement.

**Supplementary Fig. S8 | Kidney compartment cross-disease responsiveness.** Supporting the kidney vignette: ranking of seven functional compartments by adjusted effect size across non-renal prevalent and incident diseases; kidney age gap vs. HLS association; kidney disease lifestyle-associated risk offset compared to other endpoints. Referenced in Discussion: lifestyle and compartment-level interpretation.

**Supplementary Fig. S9 | Longitudinal validation of OAG.** Two-panel figure (N = 4,789 with repeat MRI). a, Cross-sectional HLS–OAG replication at Visit 3 (β = −0.132, P = 1.4 × 10⁻⁴). b, ΔHLS vs. ΔOAG (null: β = −0.003, P = 0.93). Referenced in Discussion: Lifestyle paragraph and Limitations.

**Supplementary Fig. S10 | Lifestyle habit transitions between Visit 2 and Visit 3.** Domain-specific transition rates for smoking, physical activity, diet, alcohol and sleep in the repeat-imaging subset (N = 4,764). Referenced in Discussion: Lifestyle paragraph and Limitations.

**Supplementary Fig. S11 | Study flow diagram.** CONSORT-style flow diagram showing participant selection from 69,546 with abdominal MRI to: N = 67,130 with successful predictions across all eight compartments; N = 56,525 primary analytical cohort for prevalent disease, biomarker and lifestyle analyses (after covariate completeness and exclusion of model development/evaluation participants, N = 7,715); N = 20,266 prospective endpoint cohort used for both incident disease and all-cause mortality analyses (after additional blanket prevalent-disease baseline exclusion); and N = 40,985 separate broader-cohort mortality sensitivity analysis (drawn from the analytical cohort using a narrower exclusion list).

**Supplementary Table S6. | Anatomical negative-control associations.** Eye and ear ICD-10 categories (anatomical structures outside the abdominal imaging field of view) were retained as anatomical negative controls. OAG was not significantly associated with any eye or ear endpoint at FDR < 0.05 across endpoints, supporting the anatomical specificity of the abdominal aging signal. Referenced in Results: prevalent disease.

## Data Availability

All data analyzed in the present study were obtained from the UK Biobank Resource (Application Number 132578) and are available to bona fide researchers via application to UK Biobank at https://www.ukbiobank.ac.uk/. The derived Overall Aging Gap and compartment-specific predicted age gap values used in the statistical analyses will be returned to UK Biobank upon publication for use by other approved researchers.

## References

1. Tian, Y. E. et al. Heterogeneous aging across multiple organ systems and prediction of chronic disease and mortality. Nat. Med. 29, 1221–1231 (2023).

2. Oh, H. S.-H. et al. Organ aging signatures in the plasma proteome track health and disease. Nature 624, 164–172 (2023).

3. Wen, J. et al. MRI-based multi-organ clocks for healthy aging and disease assessment. Nat. Med. 32, 82–92 (2026).

4. Ren, P. et al. Imaging-based organ-specific aging clock predicts human diseases and mortality. npj Digit. Med. 9, 278 (2026). doi:10.1038/s41746-026-02488-7

5. Ecker, V., Früh, M., Yang, B., Gatidis, S. & Küstner, T. Deep regression for biological age estimation in multiple organs: Investigations on 40,000 subjects of the UK Biobank. In Proc. IEEE ICASSP 2255–2259 (2024).

6. Ecker, V., Yang, B., Gatidis, S. & Küstner, T. Imaging-derived biological age across multiple organs links to mortality and aging-related health outcomes. npj Aging (2026). doi:10.1038/s41514-026-00377-7

7. Zhang, R. et al. Brain age gap as a predictive biomarker that links aging, lifestyle, and neuropsychiatric health. Commun. Med. 5, 441 (2025).

8. Le Goallec, A. et al. Using deep learning to predict abdominal age from liver and pancreas magnetic resonance images. Nat. Commun. 13, 1979 (2022).

9. Cruz-Jentoft, A. J. et al. Sarcopenia: revised European consensus on definition and diagnosis. Age Ageing 48, 16–31 (2019).

10. Ou, M.-Y., Zhang, H., Tan, P.-C., Zhou, S.-B. & Li, Q.-F. Adipose tissue aging: mechanisms and therapeutic implications. Cell Death Dis. 13, 300 (2022).

11. Littlejohns, T. J. et al. The UK Biobank imaging enhancement of 100,000 participants: rationale, data collection, management and future directions. Nat. Commun. 11, 2624 (2020).

12. Roy, S. et al. MedNeXt: Transformer-driven scaling of ConvNets for medical image segmentation. In Proc. MICCAI 405–415 (2023).

13. Graf, R., et al. TotalVibeSegmentator: full body MRI segmentation for the NAKO and UK Biobank. arXiv preprint arXiv:2406.00125 (2024).

14. Kart, T. et al. Deep learning-based automated abdominal organ segmentation in the UK Biobank and German National Cohort magnetic resonance imaging studies. Invest. Radiol. 56, 401–408 (2021).

15. Argentieri, M. A. et al. Proteomic aging clock predicts mortality and risk of common age-related diseases in diverse populations. Nat. Med. 30, 2450–2460 (2024).

16. Cumplido-Mayoral, I. et al. The mediating role of neuroimaging-derived biological brain age in the association between risk factors for dementia and cognitive decline in middle-aged and older individuals without cognitive impairment: a cohort study. Lancet Healthy Longev. 5, e276–e286 (2024).

17. Kivimäki, M. et al. Proteomic organ-specific ageing signatures and 20-year risk of age-related diseases: the Whitehall II observational cohort study. Lancet Digit. Health 7, e195–e204 (2025).

18. Wang, Y., Xiao, S. et al. Organ-specific proteomic aging clocks predict disease and longevity across diverse populations. Nat. Aging 6, 162–180 (2026).

19. Denic, A., Lieske, J. C., Chakkera, H. A. et al. The substantial loss of nephrons in healthy human kidneys with aging. J. Am. Soc. Nephrol. 28, 313–320 (2017).

20. Denic, A., Glassock, R. J. & Rule, A. D. Structural and functional changes with the aging kidney. Adv. Chronic Kidney Dis. 23, 19–28 (2016).

21. Sehgal, R. et al. Systems Age: a single blood methylation test to quantify aging heterogeneity across 11 physiological systems. Nat. Aging 5, 1880–1896 (2025).

22. Huang, Y. et al. Healthy lifestyle habits, educational attainment, and the risk of 45 age-related health and mortality outcomes in the UK: a prospective cohort study. J. Nutr. Health Aging 29, 100525 (2025).

